# Using publicly available data to identify priority communities for a SARS-CoV-2 testing intervention in a southern U.S. state

**DOI:** 10.1101/2023.01.31.23285248

**Authors:** Lynn T Matthews, Dustin M Long, Madeline C Pratt, Ya Yuan, Sonya L Heath, Emily B Levitan, Sydney Grooms, Thomas Creger, Aadia Rana, Michael J Mugavero, Suzanne E Judd, the COVID COMET RADXUP Team

**Affiliations:** Division of Infectious Disease, Department of Medicine, Heersink School of Medicine, University of Alabama at Birmingham, Birmingham, Alabama, USA; Department of Biostatistics, School of Public Health, University of Alabama at Birmingham, Birmingham, Alabama, USA; Department of Epidemiology, School of Public Health, University of Alabama at Birmingham, Birmingham, Alabama, USA; Center for AIDS Research, Heersink School of Medicine, University of Alabama at Birmingham, Birmingham, Alabama, USA; Center for the Study of Community Health, School of Public Health, University of Alabama at Birmingham, Birmingham, Alabama, USA

**Keywords:** SARS-CoV-2, COVID-19 testing, population health, epidemiology, community engagement

## Abstract

**Background:** The U.S. Southeast has a high burden of SARS-CoV-2 infections and COVID-19 disease. We used public data sources and community engagement to prioritize county selections for a precision population health intervention to promote a SARS-CoV-2 testing intervention in rural Alabama during October 2020 and March 2021.

**Methods:** We modeled factors associated with county-level SARS-CoV-2 percent positivity using covariates thought to associate with SARS-CoV-2 acquisition risk, disease severity, and risk mitigation practices. Descriptive epidemiologic data were presented to scientific and community advisory boards to prioritize counties for a testing intervention.

**Results:** In October 2020, SARS-CoV-2 percent positivity was not associated with any modeled factors. In March 2021, premature death rate (aRR 1.16, 95% CI 1.07, 1.25), percent Black residents (aRR 1.00, 95% CI 1.00, 1.01), preventable hospitalizations (aRR 1.03, 95% CI 1.00, 1.06), and proportion of smokers (aRR 0.231, 95% CI 0.10, 0.55) were associated with average SARS-CoV-2 percent positivity. We then ranked counties based on percent positivity, case fatality, case rates, and number of testing sites using individual variables and factor scores. Top ranking counties identified through factor analysis and univariate associations were provided to community partners who considered ongoing efforts and strength of community partnerships to promote testing to inform intervention.

**Conclusions:** The dynamic nature of SARS-CoV-2 proved challenging for a modelling approach to inform a precision population health intervention at the county level. Epidemiological data allowed for engagement of community stakeholders implementing testing. As data sources and analytic capacities expand, engaging communities in data interpretation is vital to address diseases locally.

## Introduction

From the outset of the global pandemic, the burden of COVID-19 disease has been dynamic across person, time, and space. In the months of the U.S. pandemic prior to vaccine availability, SARS-CoV-2 testing demand outweighed supply[1]. Further, those most vulnerable to COVID-19 disease were not the most likely to be tested, and states with larger numbers of people living in poverty and with poorer infrastructure for public health and workers’ rights struggled to roll out testing and contact tracing [2].

Alabama is one of the lowest ranked states in many health metrics compared to the rest of the U.S. [3]. Social determinants of health (SDH) including socioeconomic status, educational attainment, racial discrimination, and restrictive governmental policies underlie these poor outcomes [4], with rural regions and Black communities disproportionately impacted [5-7]. Statewide geographic, socioeconomic, and racial disparities are reflected in how Alabama experiences the COVID-19 pandemic. In mid-July 2021, prior to the start of the SARS-CoV-2 delta strain surge, an estimated 11% of Alabama’s nearly five million people had tested for SARS-CoV-2, with approximately 4,500 tests conducted daily, or 148 per 100,000 people. The case rate per 100,000 in some rural counties was nearly triple the rate of metropolitan areas such as Mobile and Birmingham, with SARS-CoV-2 testing percent positivity approaching 20% in a majority of rural counties in July. While 27% of Alabama’s population is Black, nearly 45% of the state’s lab-confirmed SARS-CoV-2 cases and deaths are among Black people [8, 9]. Grounded in a legacy of slavery manifested in institutionalized racism [10], contemporary barriers to health include social and structural factors that heighten risk for COVID-19 disease among Black communities, particularly those in rural areas [11].

In Alabama and similar settings, there is a need to optimize benefits of testing without exacerbating stigma and marginalization in underserved communities in order to control disease spread. We were funded to collaborate with academic institutions, local service organizations, and community health workers to promote SARS-CoV-2 testing in underserved communities in Alabama. We planned to use publicly available data on SARS-CoV-2, including risk mitigation practices and vulnerability to SARS-CoV-2 infection, to identify rural counties that would benefit most from community-based testing interventions.

Here we describe how we used public data sources to explore strategies for identifying communities most at need for SARS-CoV-2 testing in two waves of selection during October 2020 and March 2021. We present our considerations and how our approach evolved with the pandemic, and lessons learned regarding the limitations of percent positivity in a state with inconsistent testing uptake to inform future work in other more rural settings with poorer public health infrastructure and healthcare.

## Methods

### Setting

The Alabama Department of Public Health announced the first SARS-CoV-2 positive test result on March 13, 2020 [12]. As of September 7, 2021, nearly 18 months later, there have been a total of 727,360 cases and 12,420 deaths in Alabama [13] and about 39% of the population is vaccinated (2 doses) [13, 14]. Alabama’s population is approximately 69% White, 27% Black, with 86% high school graduates, and approximately 16% live in poverty [15]. For reference, the U.S. census estimates that the nation’s population is approximately 76% White, 14% Black, with 89% high school graduates, and approximately 12% live in poverty [16].

### SARS-CoV-2 testing data

Data were downloaded periodically (approximately quarterly) from publicly available websites that collated and created visual representations of data reported by the Alabama Department of Public Health (ADPH). For the October 2020 county testing selections, data were downloaded from bamatracker.com[17], a website that collated and displayed data from ADPH through May 2021. In the March 2021 round of selections, data were downloaded from bamatracker.com[13] as well as the New York Times website[18].

### Summary models reviewed

We reviewed existing models summarizing, modeling, and predicting patterns in SARS-CoV-2 testing data provided by Johns Hopkins University[19], University of Washington (Institute for Health Metrics and Evaluation, IHME)[20], and the Pandemic Vulnerability Index (PVI) from National Institute of Environmental Health Sciences [21]. We reviewed the COVID Health Equity interactive dashboard summarized by Emory University[22]. None of these were able to distinguish testing intervention need in Alabama at the county level as the entire state was in highest risk category for COVID-19 disease with widespread need for increased SARS-CoV-2 testing in both rounds of selections. However, elements of data sources used in these models and collated on their websites were incorporated as described in model covariates below.

### Model building using local data

#### Outcome

Percent positivity, or the number of positive tests per 100 tests performed, has been widely used as a measure of SARS-CoV-2 disease burden since the beginning of the pandemic, providing insights into transmission within specific geographic areas[23]. The CDC recommends percent positivity as a measure of disease surveillance for public health decision-making[23]. We modeled factors associated with SARS-CoV-2 percent positivity with the goal to identify factors associated with higher percent positivity to inform intervention county selections. At the time of these selections, most Alabama counties reported high percent positivity rates and were uniformly identified as highest risk by the available models. We therefore intended to identify factors that drove that risk and focus selections on counties based on the distributions of factors associated with higher SARS-CoV-2 percent positivity. Our models differed slightly with the evolution in the pandemic, knowledge of SARS-CoV-2 risk factors, and features of the ADPH testing data between October 2020 and March 2021.

#### October 2020

We used linear regression models with dispersion with the primary outcome of average 14-day SARS-CoV-2 percent positivity summarized at the county level from October 2 – October 16, 2020. Model covariates included factors associated with risk of acquiring SARS-CoV-2 (Daytime Population Density [21, 24], SVI Housing Score [4, 21], % Below Poverty Line [24, 25], % Unemployed [24], % Black [21], % High School graduates [24, 25], % Limited English [24, 25], COVID Testing Sites [13], increased risk of COVID-19 disease severity (Air pollution [21, 26], Premature Death [24], % Smoking [21, 27], % Over 65 [21], % Obese [21, 27], % Diabetic [21, 28], mortality due to influenza and/or pneumonia [25, 29, 30], Number Preventable Hospitalization [25, 31, 32], COPD Prevalence [33], Heart Disease Prevalence [33]), and factors associated with mitigation behavior (% Republican voters [34]).

#### March 2021

We used negative binomial regression modeling with dispersion to evaluate factors associated with average 7-day SARS-CoV-2 percent positivity at the county level from 06/17/2020 -3/31/2021. Between October and March, we learned that for some months, the 14-day percent positivity result produced in the state data eliminated all individuals with prior SARS-CoV-2 testing data from the denominator, thus inflating the 14-day percent positivity. This did not occur with the 7-day summary value. Model covariates included factors associated with increased risk of acquiring SARS-CoV-2, increased risk of COVID-19 disease severity, and factors associated with less risk mitigation behavior as above.

#### Alternative Approach Using Descriptive Epidemiology

In October 2020, we summarized past 14-day average SARS-CoV-2 percent positivity, 14-day case fatality, and 14-day cases/100k population. We also included a cross sectional description of the number of testing sites per county based on collated data provided by ADPH. We described the counties in the top quartile for percent positivity, case fatality, and case rates and below the median for number of testing sites. We also conducted a factor analysis including 3-month percent SARS-CoV-2 positivity, SARS-CoV-2 case fatality rate, COVID-testing sites per 100k population. One factor explained 41% of the variability and was used to provide an additional ranking of counties for selection.

In March 2021, we summarized first quarter 2021 (01/01/2021-03/31/2021) 90-day average case fatality, case rate, and overall mortality rate by county. We identified the counties in the top quartile for percent positivity, case fatality, case rates, and mortality.

In both rounds of selections, we also described race[24, 25], poverty[24, 25], and rurality[21, 24] to inform selections. In the March 2021 selections, we also described vaccine uptake, reported by ADPH.

#### Preliminary county selections

For both the October 2020 and March 2021 selections, the counties with the greatest COVID-19 burden as described above were shared with our Scientific and Community Advisory Boards to further hone selections and avoid overlapping outreach with other efforts (e.g., ADPH focus areas, other RADX-Up projects in the region) and to prioritize counties in need with known partners to promote community outreach efforts. Final selections were made by the Scientific and Community Advisory Boards in collaboration with the scientific leadership for this project.

## Results

In October 2020, we modeled the predictors of past 14-day percent SARS-CoV-2 positivity and most recent county-level estimates for clinical factors, social determinants of health, and other measures of behavioral risk mitigation strategies we expected to inform testing and case rates. No covariates were significantly associated with county level percent positivity (Table 1).

**Table 1.**
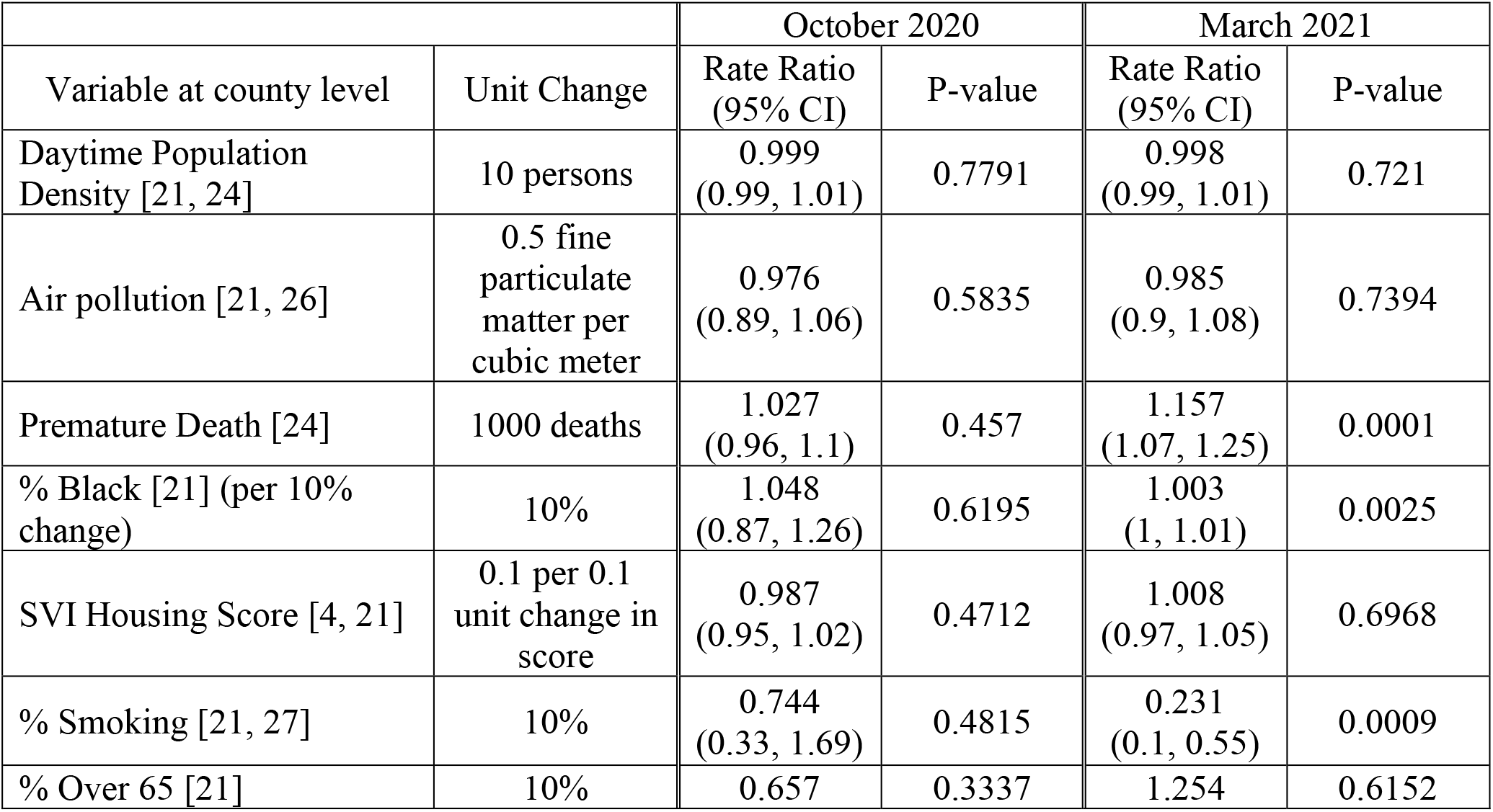
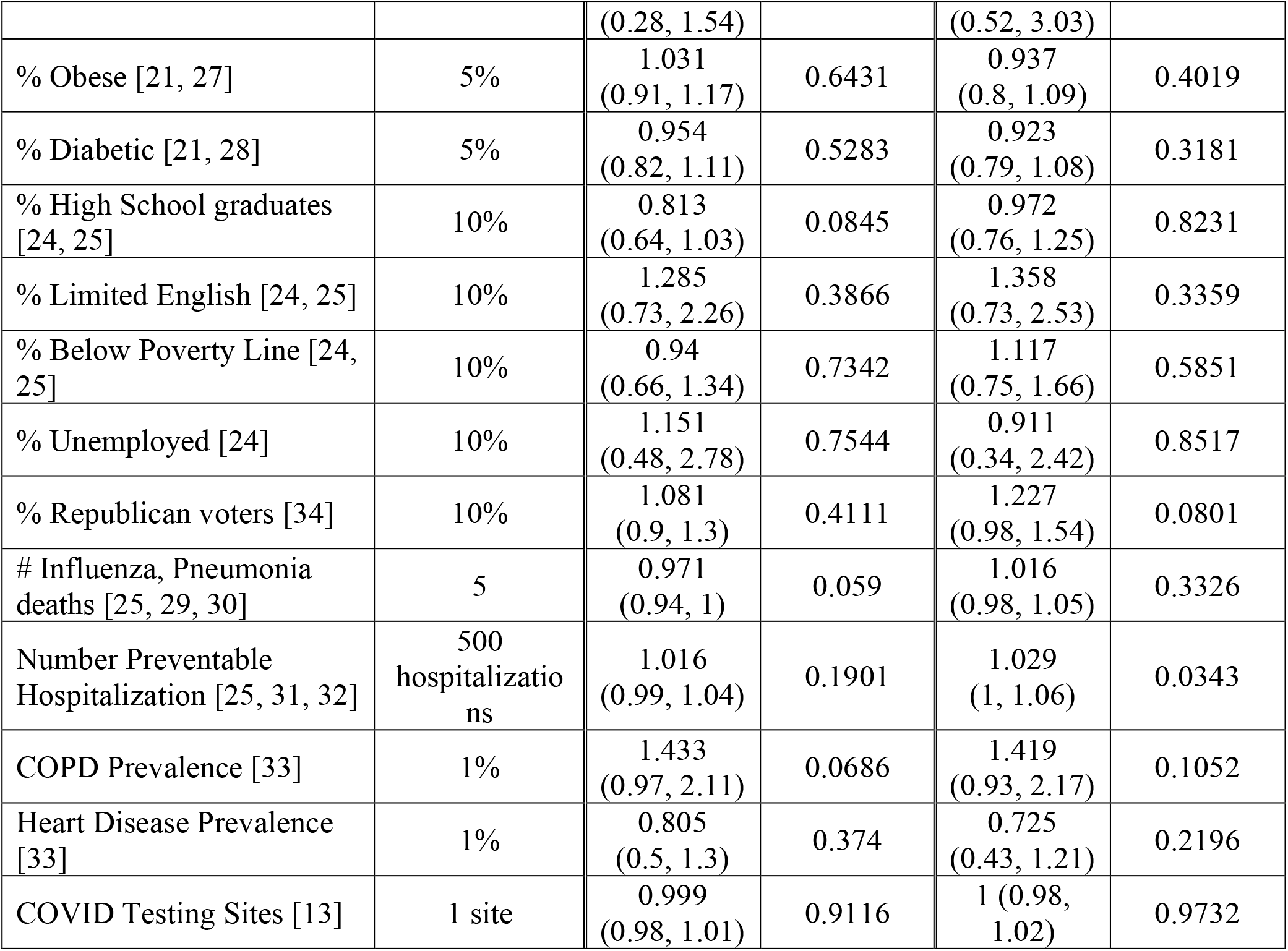
Model outputs for SARS-CoV-2 percent positivity at two timepoints in Alabama’s 67 counties.

In March 2021, we modeled predictors of average 7-day case positivity from 6/18/2020 – 3/31/2021 using similar covariates (Table 1). In this adjusted model, premature death rate was associated with 16% higher SARS-CoV-2 percent positivity (aRR 1.16, 95% CI 1.07, 1.25) per 1,000 deaths, 10% greater proportion of county residents identifying as Black was associated with a <1% higher in SARS-CoV-2 percent positivity (aRR 1.003, 95% CI 1.00, 1.01), and number of preventable hospitalizations was associated with a slightly higher in SARS-CoV-2 percent positivity (aRR 1.03, 95% CI 1.00, 1.06) per 500 hospitalizations). A 10% higher proportion of the county population identifying as smokers was associated with a 77% lower percent positivity (aRR 0.231, 95% CI 0.10, 0.55).

Given the limited predictive power of the existing models and the models we built to identify counties with the greatest need, our county selections were determined based on descriptive epidemiology and input from scientific and community advisory boards. We described the counties in the top quartile for percent positivity, case fatality and case rates and below the median for number of testing sites and organized counties based on the number of parameters for which they were in the top quarter of the 67 counties in the state (Table 2). Table 3 demonstrates the October 2020 summary of past 14-day SARS-CoV-2 percent positivity, 14-day case fatality, and 14-day cases/100k population. We included a cross sectional description of the number of testing sites per county based on collated data provided by ADPH.

**Table 2.** Descriptive epidemiology used to inform October 2020 county level selections for a SARS-CoV-2 testing intervention in Alabama.

| County | Testing sites/100k | 3M % Positivity | 3M # cases/100k | 14D % pos | 14D # Cases/100k | Case fatality | % AA | % Hispanic | Population | % poverty | Preventable Hospitalizations Rate per 100K | Ranking by marker of HC system quality (1=highest) |
| --- | --- | --- | --- | --- | --- | --- | --- | --- | --- | --- | --- | --- |
| Franklin | 3.2 | 18.49 | 6431.14 | 26.50 | 436.82 | 1.63 | 3.88 | 16.9 | 31363 | 14.80 | 7810 | 5 |
| Clarke | 4.2 | 13.51 | 5321.91 | 17.81 | 510.03 | 1.41 | 44.33 | 0.2 | 23920 | 18.40 | 5857 | 30 |
| Clay | 7.5 | 20.33 | 5438.79 | 27.00 | 610.17 | 1.99 | 13.83 | 3.1 | 13275 | 14.80 | 5644 | 35 |
| Lawrence | 3.0 | 16.96 | 2460.78 | 22.85 | 379.28 | 4.04 | 10.83 | 2.1 | 32957 | 13.40 | 6598 | 14 |
| Dallas | 7.8 | 10.37 | 4855.13 | 10.09 | 120.07 | 1.67 | 70.06 | 1 | 38310 | 24.80 | 6574 | 16 |
| Conecuh | 16.3 | 14.99 | 4528.79 | 13.08 | 114.03 | 2.77 | 45.66 | 0.8 | 12277 | 17.40 | 11309 | 2 |
| Russell | 3.5 | 12.97 | 3296.93 | 12.89 | 169.61 | 0.17 | 44.76 | 5.4 | 57781 | 15.40 | 4027 | 63 |
| Montgomery | 5.3 | 13.09 | 4349.69 | 16.96 | 265.32 | 2.09 | 58.55 | 3.4 | 225763 | 16.30 | 5809 | 32 |
| Lowndes | 10.0 | 14.71 | 7038.3 | 26.40 | 330.86 | 3.77 | 71.94 | 0.7 | 9974 | 23.40 | 5337 | 43 |
| DeKalb | 8.4 | 16.05 | 4558.38 | 31.42 | 745.25 | 0.88 | 1.45 | 14.4 | 71385 | 17.00 | 5204 | 45 |
| Washington | 6.1 | 13.03 | 3803.88 | 19.12 | 317.50 | 2.12 | 23.33 | 1.1 | 16378 | 16.30 | 4939 | 49 |
| Limestone | 3.1 | 14.05 | 2844.84 | 26.25 | 421.11 | 1.02 | 13.49 | 5.9 | 96174 | 10.20 | 5951 | 27 |
| Chilton | 9.1 | 17.20 | 4135.62 | 17.17 | 332.93 | 1.71 | 9.97 | 7.8 | 44153 | 15.30 | 3791 | 66 |
| Morgan | 2.5 | 12.63 | 3358.83 | 21.72 | 389.62 | 0.84 | 12.66 | 8.1 | 119089 | 12.50 | 5862 | 29 |
| Randolph | 4.4 | 15.39 | 3542.35 | 26.27 | 409.24 | 1.81 | 19.36 | 2.9 | 22725 | 14.30 | 4215 | 61 |
| Lamar | 7.2 | 13.52 | 3315.52 | 25.73 | 382.84 | 0.75 | 10.39 | 1.6 | 13844 | 15.20 | 5852 | 31 |
| Marshall | 4.2 | 13.12 | 4484.49 | 15.58 | 221.62 | 1.22 | 2.40 | 13.6 | 96109 | 16.20 | 5877 | 28 |

Darkest shaded counties met 4 of 4 criteria, next darkest met 3 of 4, lighter gray met 2 of 4. The criteria (columns in box) were top quartile for (1) case rate, (2) percent positivity, (3) case fatality, and (4) death rate.

**Table 3.** Top 10 counties based on Factor Analysis including case fatality rate, case positivity, SARS-CoV-2 testing sites per 100K population for the October 2020 selections.

| County | Factor Score | 3-month SARS-CoV-2 % Positivity | Case fatality rate | SARS-CoV-2 Testing Sites per 100K population |
| --- | --- | --- | --- | --- |
| Franklin | -1.80 | 18.49 | 1.63 | 3.19 |
| Clay | -1.74 | 20.34 | 1.99 | 7.53 |
| Russell | -1.37 | 12.97 | 0.17 | 3.46 |
| DeKalb | -1.36 | 16.05 | 0.88 | 8.41 |
| Limestone | -1.22 | 14.05 | 1.02 | 3.12 |
| Chilton | -1.16 | 17.20 | 1.71 | 9.06 |
| Morgan | -1.05 | 12.63 | 0.84 | 2.52 |
| Randolph | -1.04 | 15.39 | 1.81 | 4.40 |
| Lamar | -0.99 | 13.52 | 0.75 | 7.22 |
| Marshall | -0.88 | 13.12 | 1.22 | 4.16 |

Factor scores provided a ranking of counties with respect to 3 months SARS-CoV-2 positivity, case fatality rate, and SARS-CoV-2 testing sites/100k population. The factor rankings indicated in Table 3 can be interpreted as how much they differ from the average county on the composite measure of SARS-CoV-2 test positivity, case fatality, and testing sites. For example, Franklin County was estimated to be about 1.8 standard deviations worse compared to the average. The factor analysis ranking is described in Table 3 and identified the top 10 counties with the highest ranking based on the combined factor analysis.

Fig 1 indicates which counties were prioritized for the intervention when considering the overlapping counties identified by the factor analysis and the descriptive epidemiology and with consideration of strensgth of partnerships and other ongoing efforts to promote testing across the state.

**Figure 1.**
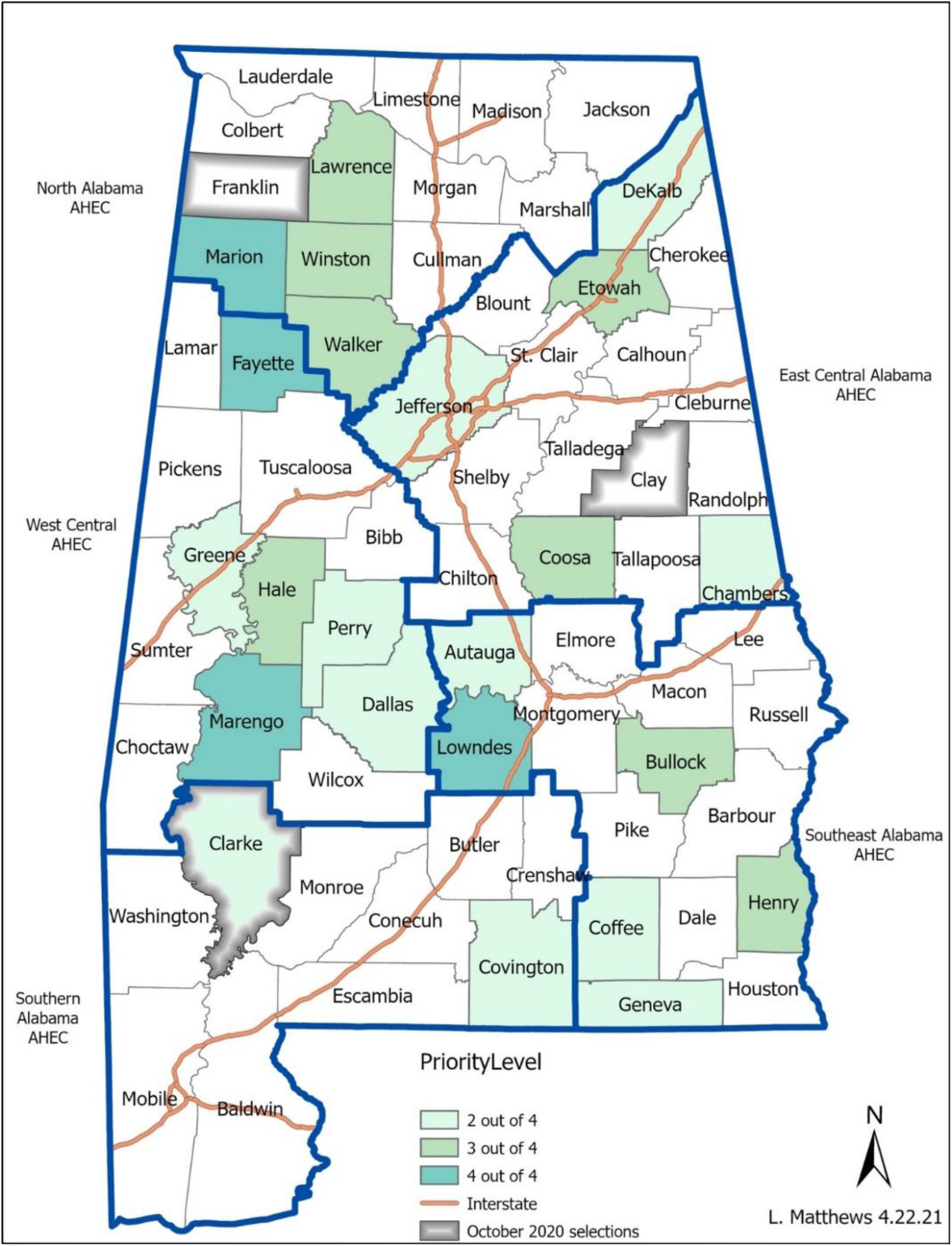
Priority county selections.

Priority counties selected in both phases of the project based on the descriptive epidemiology and mapped by the service areas of our community partners (Area Health Education Centers, AHEC) who conducted the testing.

Given the ongoing lack of discrimination between counties in the models we again used descriptive epidemiology and input from scientific and community advisory boards for the March 2021 selections. Table 4 demonstrates the March 2021 summaries of first quarter 2021 (01/01/2021-03/31/2021) 90-day average case fatality, case rate, and overall mortality rate by county. We identified the counties in the top quartile for percent positivity, case fatality, case rates, and mortality. Fig 1 indicates the counties prioritized for the intervention when considering the descriptive epidemiology, strength of partnerships, and other ongoing efforts to promote testing across the state.

**Table 4.**
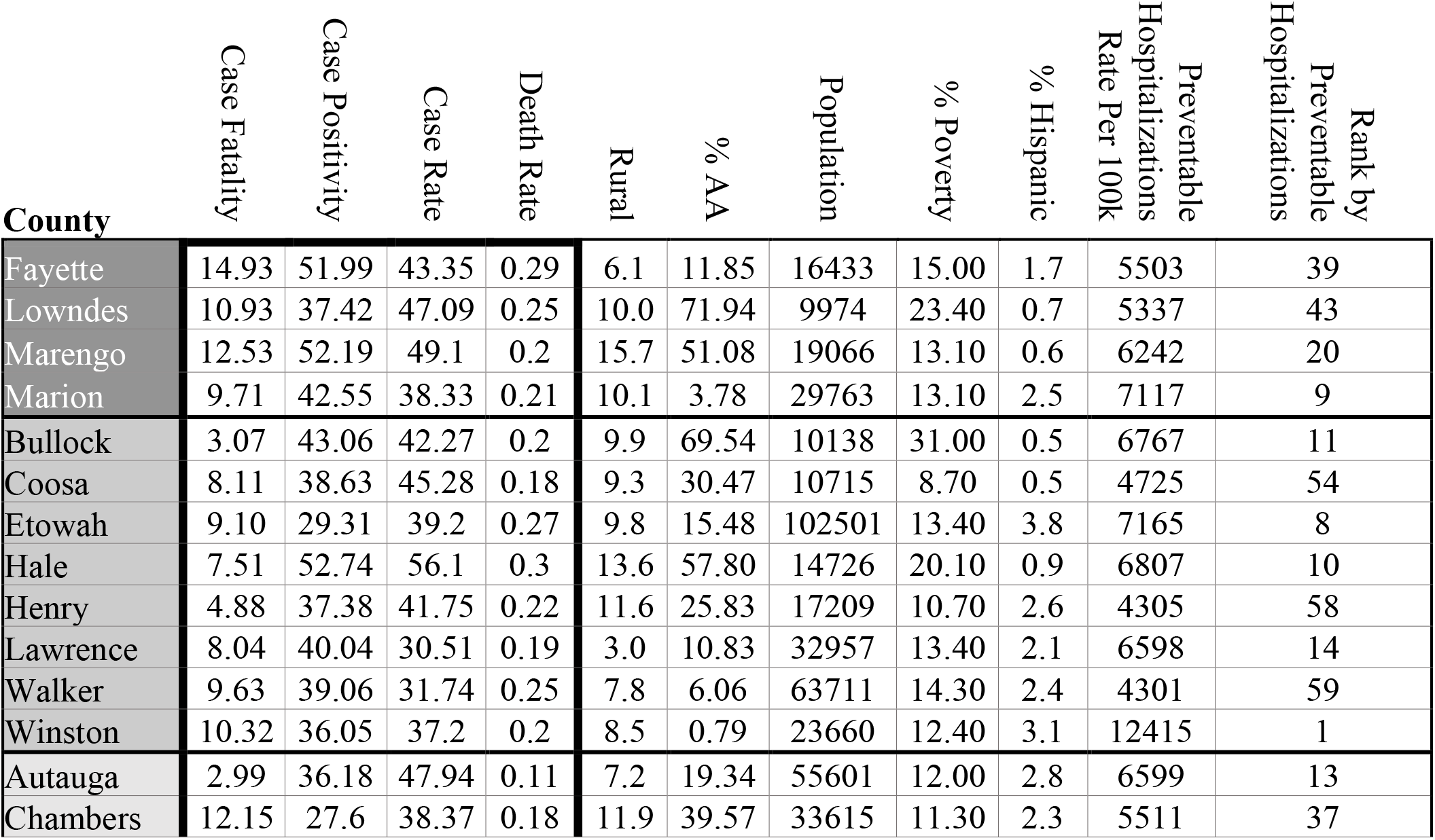
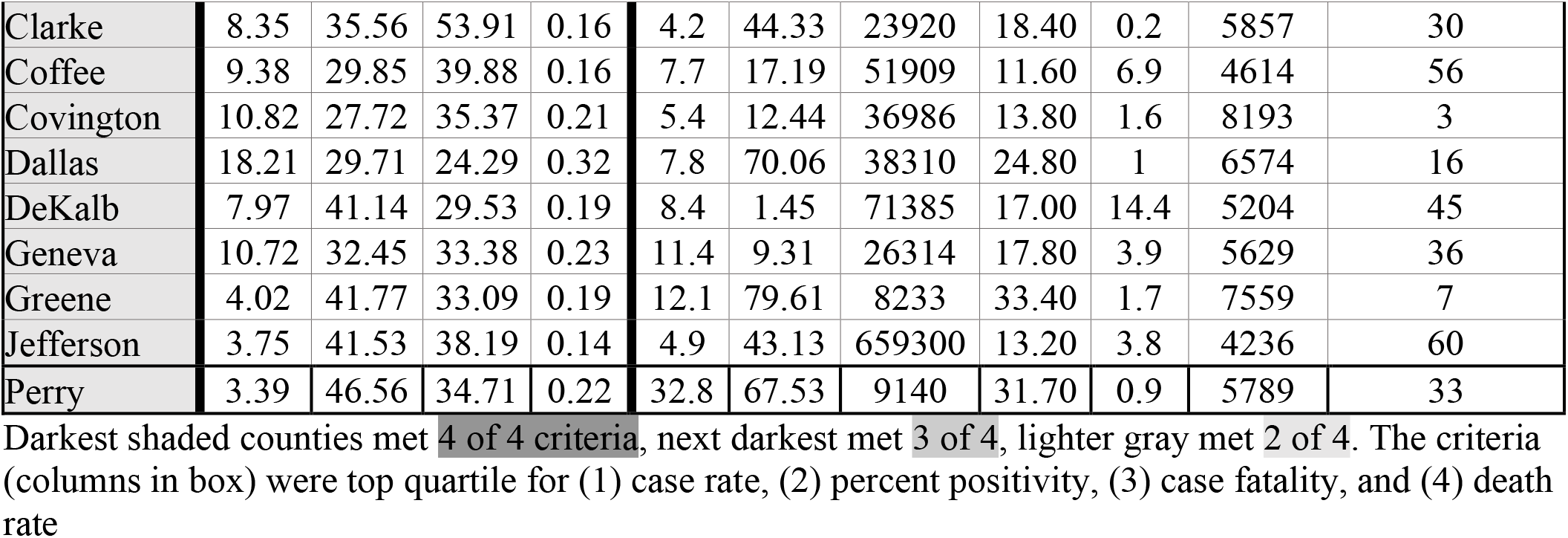
Descriptive epidemiology used to inform March 2021 county level selections for a SARS-CoV2 testing intervention in Alabama.

| County | Case Fatality | Case Positivity | Case Rate | Death Rate | Rural | % AA | Population | % Poverty | % Hispanic | Preventable Hospitalizations Rate Per 100k | Rank by Preventable Hospitalizations |
| --- | --- | --- | --- | --- | --- | --- | --- | --- | --- | --- | --- |
| Fayette | 14.93 | 51.99 | 43.35 | 0.29 | 6.1 | 11.85 | 16433 | 15.00 | 1.7 | 5503 | 39 |
| Lowndes | 10.93 | 37.42 | 47.09 | 0.25 | 10.0 | 71.94 | 9974 | 23.40 | 0.7 | 5337 | 43 |
| Marengo | 12.53 | 52.19 | 49.1 | 0.2 | 15.7 | 51.08 | 19066 | 13.10 | 0.6 | 6242 | 20 |
| Marion | 9.71 | 42.55 | 38.33 | 0.21 | 10.1 | 3.78 | 29763 | 13.10 | 2.5 | 7117 | 9 |
| Bullock | 3.07 | 43.06 | 42.27 | 0.2 | 9.9 | 69.54 | 10138 | 31.00 | 0.5 | 6767 | 11 |
| Coosa | 8.11 | 38.63 | 45.28 | 0.18 | 9.3 | 30.47 | 10715 | 8.70 | 0.5 | 4725 | 54 |
| Etowah | 9.10 | 29.31 | 39.2 | 0.27 | 9.8 | 15.48 | 102501 | 13.40 | 3.8 | 7165 | 8 |
| Hale | 7.51 | 52.74 | 56.1 | 0.3 | 13.6 | 57.80 | 14726 | 20.10 | 0.9 | 6807 | 10 |
| Henry | 4.88 | 37.38 | 41.75 | 0.22 | 11.6 | 25.83 | 17209 | 10.70 | 2.6 | 4305 | 58 |
| Lawrence | 8.04 | 40.04 | 30.51 | 0.19 | 3.0 | 10.83 | 32957 | 13.40 | 2.1 | 6598 | 14 |
| Walker | 9.63 | 39.06 | 31.74 | 0.25 | 7.8 | 6.06 | 63711 | 14.30 | 2.4 | 4301 | 59 |
| Winston | 10.32 | 36.05 | 37.2 | 0.2 | 8.5 | 0.79 | 23660 | 12.40 | 3.1 | 12415 | 1 |
| Autauga | 2.99 | 36.18 | 47.94 | 0.11 | 7.2 | 19.34 | 55601 | 12.00 | 2.8 | 6599 | 13 |
| Chambers | 12.15 | 27.6 | 38.37 | 0.18 | 11.9 | 39.57 | 33615 | 11.30 | 2.3 | 5511 | 37 |
| Clarke | 8.35 | 35.56 | 53.91 | 0.16 | 4.2 | 44.33 | 23920 | 18.40 | 0.2 | 5857 | 30 |
| Coffee | 9.38 | 29.85 | 39.88 | 0.16 | 7.7 | 17.19 | 51909 | 11.60 | 6.9 | 4614 | 56 |
| Covington | 10.82 | 27.72 | 35.37 | 0.21 | 5.4 | 12.44 | 36986 | 13.80 | 1.6 | 8193 | 3 |
| Dallas | 18.21 | 29.71 | 24.29 | 0.32 | 7.8 | 70.06 | 38310 | 24.80 | 1 | 6574 | 16 |
| DeKalb | 7.97 | 41.14 | 29.53 | 0.19 | 8.4 | 1.45 | 71385 | 17.00 | 14.4 | 5204 | 45 |
| Geneva | 10.72 | 32.45 | 33.38 | 0.23 | 11.4 | 9.31 | 26314 | 17.80 | 3.9 | 5629 | 36 |
| Greene | 4.02 | 41.77 | 33.09 | 0.19 | 12.1 | 79.61 | 8233 | 33.40 | 1.7 | 7559 | 7 |
| Jefferson | 3.75 | 41.53 | 38.19 | 0.14 | 4.9 | 43.13 | 659300 | 13.20 | 3.8 | 4236 | 60 |
| Perry | 3.39 | 46.56 | 34.71 | 0.22 | 32.8 | 67.53 | 9140 | 31.70 | 0.9 | 5789 | 33 |

Darkest shaded counties met 4 of 4 criteria, next darkest met 3 of 4, lighter gray met 2 of 4. The criteria (columns in box) were top quartile for (1) case rate, (2) percent positivity, (3) case fatality, and (4) death rate

Fig 1 highlights the priority counties selected in phase 1 in gray. This figure demonstrates how candidate counties (shades of green) were presented to the community and scientific advisory boards for the March 2021 selections, including data on the regions served by the service areas of our community partners (Area Health Education Centers, AHEC) who conducted the testing, in blue. For the March 2021 phase, the final decision was to intervene on regions rather than counties given widespread need and the nature of our community partnerships.

## Discussion

Here we present our approach to pragmatic identification of rural counties in need of a SARS-CoV-2 testing intervention in a Southern U.S. state at the peak of the 2019-first quarter 2021 phase of the U.S. SARS-CoV-2 epidemic. In order to select counties for rapid deployment of a testing intervention in a state with high testing need, we were tasked with quickly identifying priority counties using publicly-available data. Based on global and national guidance, we focused on SARS-CoV-2 percent positivity[17, 22]. As models evaluating factors associated with percent positivity did not discriminate counties for selection, we pivoted to analyses of descriptive epidemiological data to inform county selection. This approach allowed us to categorize counties based upon the count of four severity criteria with four and 12 of Alabama’s 68 counties meeting at least three of these criteria in October 2020 and March 2021, respectively. The presentation of these data in tabular and geospatial fashion was easily interpreted by the community and scientific advisory boards, and community testing implementation partners, with prioritization of counties for testing further benefitting from an understanding of the local community context in identified high-severity counties. Equipped with this information, as depicted in Fig 1, community testing partners were able to prioritize rural counties for outreach and the identification of venues and local partners for testing delivery [35]. As our data sources and analytic capacities expand in scope and sophistication, leveraging traditional epidemiological data and analysis retains a vital role in engaging communities to address communicable and non-communicable conditions in their local context, particularly when response to emerging diseases when time for sophisticated analyses is limited.

In October 2020, no factors were associated with percent positivity in our models. This speaks to the fact that in these first waves of COVID-19 no population was immune, and testing was roughly similar across the counties, so it is not surprising that specific factors driving COVID transmission and positivity were not identified. In addition, more localized outbreaks and “super-spreader” events were randomly distributed in time and geographic area and therefore the time periods included in the models represent variable features of a dynamic epidemic. In March 2021, factors associated with increases in percent positivity were county level rates of premature death[24] and preventable hospitalizations[25, 31, 32]. In addition, having a higher proportion of the population who identified as Black increased the likelihood of case positivity. These associations may reflect that those in poorer resourced counties were at higher risk of acquiring SARS-CoV-2 and also less likely to have access to or seek testing in the absence of infection. In addition, an increase in the proportion of county residents who self-described as smokers was associated with a decrease in the SARS-CoV-2 percent positivity. This may reflect more screening on the part of smokers and those living with smokers who are at higher risk for respiratory disease, in general, and possibly for SARS-CoV-2 complications [36-38]. However, there are limits to this model given county level population-level associations which are broad and subject to ecological fallacy. In both selection rounds, the models were not helpful in informing key areas to focus outreach efforts and therefore, we pivoted to descriptive epidemiology to identify areas most in need of testing with the assumption that the testing intervention might improve testing, contact tracing and thus reduce disease spread, case fatality and morbidity.

Identifying the appropriate metric to track in order to identify areas of testing demand during a rapidly evolving pandemic proved to be challenging. Case positivity was originally thought to be the best indicator of the next outbreak which would therefore inform aggressive testing strategies[39]. This did not prove to be the case due to the often-asymptomatic nature of SARS-CoV-2 and the overwhelming task of contract tracing in our strained public health setting [40, 41]. In addition, (a) errors in reporting, (b) state and institutional guidance to not test if household contact has tested positive at times when testing capacity was limited, (c) workers unwilling to test and be forced to quarantine and lose pay, (d) poorer health resourced state where people are anxious about potential test-related fees, (e) erratic approach to testing where some institutions and organizations required weekly tests for workers, (f) variations in data with large numbers of tests in Summer and Fall 2020 with push resumption of in-person university activities in a state with over 300,000[42] university students[43] changed the meaning of case positivity.

This team is led by HIV researchers who routinely wait years for public health data to be cleaned and available, in part due to the sensitive nature of HIV testing and related data [44, 45]. The excitement of real-time data releases that are key to managing the COVID-19 pandemic was tempered by frustrations when rapid data availability was associated with less reliable data. For instance, Alabama datasets, like many other datasets, had noise related to large data dumps due to institutions reporting testing data en masse (e.g. 2352 new cases reported on 3 March 2021 reflecting data from months prior) changing denominators, and, at times, incomplete data. Our process highlights the need to proceed with caution and engage with community stakeholders when using real-time, inherently messy data to inform public health interventions.

In recent years there has been a push for precision population health, leveraging big data to identify those at greatest risk to optimize delivery of the right intervention, to the right population, at the right time. Unlike diseases with more stable epidemiological and risk profiles, the dynamic nature of SARS-CoV-2, over geography and time, proved challenging for a model-based approach to inform precision population health tailored SARS-CoV-2 testing. A simplified approach based upon descriptive epidemiological data proved informative while allowing for meaningful engagement of essential community stakeholders and implementation partners ultimately charged with implementing testing in their local communities. As data sources and analytic capacities expand in scope and sophistication, leveraging traditional epidemiological data and analysis retains a vital role in engaging communities and building trust in data in order to optimally address communicable and non-communicable conditions in their local context.

## Limitations

This is a methods paper to share lessons learned in selecting counties for a SARS-CoV-2 testing intervention but not an in-depth exploration of SARS-CoV-2 testing patterns and epidemiology in Alabama. The findings are unlikely to generalize to all settings and will be most applicable to other counties or states with strained public health systems in the U.S. healthcare context. Details regarding our testing intervention and findings are published elsewhere [35].

## Conclusions

We present a pragmatic approach to inform an evolving, pandemic community-level intervention. We also share lessons learned regarding the limits of test positivity in an outbreak where testing uptake is poor and where case positivity rates generally exceeded a threshold level to discern need in areas where nearly all counties had high need. Our current and future work highlights the importance of community partnerships, local knowledge to inform testing outreach and perhaps highlights the need for more centralized pandemic preparedness.

## Data Availability

Data were downloaded periodically (approximately quarterly) from publicly available websites that collated and created visual representations of data reported by the Alabama Department of Public Health (ADPH). For the October 2020 county testing selections, data were downloaded from bamatracker.com, a website that collated and displayed data from ADPH through May 2021. In the March 2021 round of selections, data were downloaded from bamatracker.com as well as the New York Times website.

## Acknowledgements

Community partners.

Larger RADxUP team:

Greer Burkholder, MD, MSPH ^1,2,3^, Andrea Cherrington, MD, MPH ^2,3^, William Curry, MD ^2^, Latesha Elopre, MD, MSPH ^1,2^, Faith Fletcher, PhD, MA ^4^, Eric Ford, PhD, MHHA ^3,5^, Allyson Hall, PhD ^2,3,6^, Larry Hearld, PhD ^2,6^, Bertha Hidalgo, PhD, MPH ^1,2,3^, Dione King, PhD, MSW ^1,3,7^, Emily Levitan, PhD, MS ^1,2,3^, Max Michael, MD ^3^, Trisha Parekh, DO ^2^, Donna Porter, PhD ^1,2^, David Redden, PhD, MS ^3^, Michael Saag, MD ^2^, Bisaka “Pia” Sen, PhD, MA ^2,3,6^, Barbara Van Der Pol, PhD, MPH ^2^, Jeffery Walker, PhD, MA ^3,7^

COVID COMET RADXUP Team Affiliations:

1. University of Alabama at Birmingham, Center for AIDS Research, School of Medicine, Birmingham, Alabama, USA.
2. University of Alabama at Birmingham, School of Medicine, Birmingham, Alabama, USA.
3. University of Alabama at Birmingham, School of Public Health, Birmingham, Alabama, USA.
4. Baylor University, College of Medicine, Waco, Texas, USA.
5. University of Alabama at Birmingham, School of Business, Birmingham, Alabama, USA.
6. University of Alabama at Birmingham, School of Health Professions, Birmingham, Alabama, USA.
7. University of Alabama at Birmingham, College of Arts and Sciences, Birmingham, Alabama, USA.

